# Energy contribution from ultra-processed foods in Peruvian children: Frequency, Bimodality and Determinants

**DOI:** 10.1101/2024.05.24.24307617

**Authors:** Marianella Miranda-Cuadros, Miguel Campos-Sánchez, Gustavo Cediel, María Laura da Costa Louzada, Joaquín Alejandro Marrón-Ponce

## Abstract

**Objective:** To assess the energy contribution of ultra-processed foods (UPF) and its association to sociodemographic and anthropometric factors in 6-35 months old children in the databases of national surveys carried out in 2008-2010, 2015-2016 and 2019.

**Methods:** All surveys were stratified multi-stage random samples. Sub-samples had 24-hr food recall on a random day for each child, using a modified multiple step method with visual aids and scales. UPF were defined based on the Nova 4 classification system. Factors were evaluated with two generalized linear models (binomial for the percentage of UPF consumers and normal for the mean UPF energy contribution among UPF consumers only). Estimations and models were adjusted by the sampling design.

**Results:** 2887 children were included. UPF contributed 29% [CI95 27 to 31] of the total energy intake. UPF contributed 31% [30 to 33%] of energy intake among UPF consumers, decreasing with age, increasing with height-for-age and showing seasonal variation. UPF were consumed by 86% [84 to 89%] of children, increasing with age and height-for-age, slightly increasing over the years and decreasing with poverty. UPF energy contribution comes mainly from milk and derivatives 19% [17 to 20] and cereals 5% [4 to 6].

**Conclusions:** The mean UPF energy contribution and the percentage of UPF consumers vary with age, height, poverty and survey year in 6-35 months old children in Peru.

## Introduction

Overweight and obesity are among current challenges to world health^1^. In Peru 2020 both comorbidities had a prevalence of 10% in children under 5 years old^2^. A very important determinant of overweight and obesity is the intake of ultra-processed foods (UPF)^11,12^. Quoting WHO, UPF are “industrial formulations manufactured mostly or entirely from substances derived from constituents of foods, together with additives used to imitate and intensify the sensory qualities of unprocessed or minimally processed foods, and dishes and meals made from them together with processed culinary ingredients.”^3^. UPF provide excess sugar, sodium, trans and saturated fat to the diet^4^ as well as non-caloric sweeteners and additives, which are substances without nutritional value^5^.

A multinational study^6^ reported that UPF intake in 2-5 year old children was 44% of the total energy intake in Chile, 38 in Mexico, 27 in Argentina, and 18 in Colombia^7–10^. This represents a public health problem because there is some prospective evidence of association between UPF intake and adiposity in older children^11,12^.

Given the combined prevalence of overweight and obesity in Peru, this study aims to assess the energy contribution from UPF, the percentage of UPF consumers and their association with social, demographic and anthropometric factors in 6-35 month old children in Peru, using databases for two national surveys: Monitoreo Nacional de Indicadores Nutricionales (MONIN)^13^ and Vigilancia Alimentaria y Nutricional por Etapas de Vida (VIANEV)^14–16^. These results will be important for public health, since they may support strategies and policies to promote healthy eating and control overweight and obesity.

## Methods

This article presents a secondary analysis of two repeated cross-sectional national surveys (with stratified multi-stage random sampling) which evaluated children’s diet using 24-hr food recall (R24h): Monitoreo Nacional de Indicadores Nutricionales (MONIN) 2008-2010 and Vigilancia Alimentaria y Nutricional por Etapas de Vida (VIANEV) 2015-2016 and 2019.

### Population and Sample

MONIN included children under 5 years old, dividing the country in five geographic domains: Metropolitan Lima (Lima and Callao provinces), Remaining Coast (towns in the western Andean slopes below 2000 m above sea level), Urban Sierra (towns 2000 m or more above sea level, with 2000 or more inhabitants), Rural Sierra (towns 2000 m or more above sea level, with less than 2000 inhabitants) and Jungle (towns in the eastern Andean slopes below 2000 m above sea level); between 19-November-2007 and 02-April-2010.

VIANEV included children under 3 years old, dividing the country in three geographic domains: Metropolitan Lima, Urban and Rural, between October-December 2015, April-July 2016 and August-October 2019.

MONIN and VIANEV surveys had as their universe all the children residing in Peru within the stated ages and dates. The sampling design was stratified with two stages. Strata were the geographical domains further stratified by census economic level. In the first stage the sampling frame was the list of census-defined clusters as maintained by the Instituto Nacional de Estadística e Informática (INEI). From this frame, clusters were selected from each stratum by random sampling without replacement with probability proportional to the registered cluster size (about 100 households per cluster) and randomly assigned to scheduled weeks. In the second stage the eligible households in the selected clusters were enumerated by the survey teams and a simple random sample of households (10 in MONIN, 5 in VIANEV) was extracted without replacement to be interviewed. The original sample weights were calculated as the inverse of the product of the selection probabilities in both stages. This probability was re-scaled to the projected population for each year (more detail in Supplement Section S3).

For the present article inclusion criteria were: children between 6 and 35 months of age, at least one R24h measurement with energy recorded, height-for-age z-score and district identification data. From 3,022 children 6-35 months within the periods where R24h were performed, 2,887 were included.

### Food Intake Measurement

To assess the diet, the list of all foods and drinks consumed during the 24 hours before the interview was recorded using the multiple step methodology^19^ modified without the list of forgotten foods. The informant was the person in charge of the preparation and serving, being in general the child’s mother. When the child was a beneficiary of Cuna Más, a day-care nursery social program^20^, the program officers were interviewed.

The R24h was applied on a random day. In subsamples a second non-consecutive random day R24h was applied, but such data is not included in the present article.

The R24h made use of visual aids (food models, food drawings), precision scales (to weight simile foods) and updated food composition and household measures tables with equivalences for foods and meals^21,22^. Information on breast-feeding or nutritional supplements was not recorded in the R24h. The training of the interviewers was a 12-day workshop with two field pilot sessions.

### Food Classification by Industrial Processing

The criteria to categorize foods follows the Nova classification (more detail in Supplement Section S1). UPF are foods in the Nova 4 category.

Each food or drink was classified with a computer program which applied a sequence of rules to assign the Nova group depending on certain data in the consolidated food composition table (CENAN/ANDREA food group (cereals, fruits, etc.), food codes and selected words (required to be present or absent in the description)). The rules were specified by an expert nutritionist in a spreadsheet format, which was reviewed by the programmer in order to verify format requirements (spacing, case, special characters, logical expressions). The initial version of the rule files had most of the rules as wide criterion applied to food groups (for instance “all cereals but bread are Nova 2”). In the current version most of the rules are fairly specific (for instance “the word PANETON is Nova 4”).

Supplement Table S102 describes the rules by food groups. For instance, in the Cereals group we have whole grains or seeds, flour, flakes and noodles as Nova 1, cornstarch (“maicena”) as Nova 2, freshly baked bread and sugary precooked flakes as Nova 3 and bread and cereal products from several brands as Nova 4.

For labelled products not yet in the food composition tables the web sites of the food manufacturers as well as Open Food Facts were consulted.

### Covariable Measurement

The following covariables were included: age (difference between the interview date and the declared birth date), sex, anthropometry (weight-for-height and height-for-age scores), following WHO 2006 equations^17^ from the weight and height measured by the field workers, who were previously standardized for precision and accuracy^18^, geographical domain (homologated to Lima, Urban and Rural), time (year day, week day, trimester and survey year) and district poverty (proportion of households in whose income is below poverty line, as per the estimations of INEI). More detail in Supplement Section S5.

### Data Analysis

The data were recorded in paper forms during the interview for later computer data entry into a database from which anonymized analytical datasets were produced.

For this article, analytical files were consolidated homogenizing variable names and types and selecting children who fulfilled the inclusion criteria. Sampling weights were re-scaled to add up to the estimated total population for each domain.

The total energy intake and the energy contribution from each Nova category were computed for each child with consolidated food composition tables compiled from direct chemical analysis carried out continuously by CENAN ^21^ or obtained from external sources ^22^, as well as product labels collected during the surveys.

The association between UPF energy contribution with age, sex, geographical domains, survey years, anthropometric indices and poverty was evaluated with two generalized linear models (1) binomial for the percentage (/100) of UPF consumers, and (2) normal for the mean square root of the energy fraction (/1) among UPF consumers only which was transformed into energy contribution (%).

Processing and analysis were made with R 4.3.1 software and packages survey, ggplot and dplyr; all estimates, models and confidence limits were adjusted for the sampling design and weights (More details in Supplements S3 to S6).

## Results

Table 1 shows social and demographic characteristics for 2,887 children from both surveys. In the sample, 67% live in urban areas, have on average 21 months age, -0.96 height-for-age z-score, 0.45 weight-for-height z-score and 29% district poverty.

**Table 1.**

| Characteristics | Total,<br>n=2887 | MONIN, N = 723 | VIANEV, N = 2,164 |
| --- | --- | --- | --- |
| Domain, n (%) |  |  |  |
| 1:Lima | 749 (26%) | 141 (20%) | 608 (28%) |
| 2:Urban | 1,176 (41%) | 438 (61%) | 738 (34%) |
| 3:Rural | 962 (33%) | 144 (20%) | 818 (38%) |
| Sex, n (%) |  |  |  |
| female | 1,430 (50%) | 371 (51%) | 1,059 (49%) |
| male | 1,457 (50%) | 352 (49%) | 1,105 (51%) |
| Age m, mean(sd) | 20.86 (8.60) | 20.95 (8.74) | 20.83 (8.55) |
| Age Group q6m, n (%) |  |  |  |
| 6 | 591 (20%) | 152 (21%) | 439 (20%) |
| 12 | 591 (20%) | 139 (19%) | 452 (21%) |
| 18 | 577 (20%) | 142 (20%) | 435 (20%) |
| 24 | 541 (19%) | 138 (19%) | 403 (19%) |
| 30 | 587 (20%) | 152 (21%) | 435 (20%) |
| Height-for-Age z, mean(sd) | -0.96 (1.13) | -1.05 (1.20) | -0.93 (1.10) |
| Weight-for-Length z, mean(sd) | 0.45 (1.05) | 0.40 (1.06) | 0.46 (1.04) |
| Calendar Trimester, n (%) |  |  |  |
| 1 | 256 (9%) | 188 (26%) | 68 (3%) |
| 2 | 674 (23%) | 150 (21%) | 524 (24%) |
| 3 | 1,098 (38%) | 163 (23%) | 935 (43%) |
| 4 | 859 (30%) | 222 (31%) | 637 (29%) |
| District Poverty Pct., mean(sd) | 29.14 (20.38) | 35.19 (21.88) | 27.12 (19.45) |
| Total Energy kcal, mean(sd) | 995.27 (625.09) | 900.14 (585.54) | 1,027.06 (634.72) |

Unweighted. Surveys: MONIN 2008-2010, VIANEV 2015, 2016 and 2019.

Figure 1 shows the energy contribution from each Nova food category (left side) and the percentage of UPF consumers (right side), in total and subdivided by sex, age group, domain and year. Total UPF energy contribution (counting both UPF consumers and non-consumers) was 29% (CI95 27 to 30). The percentage of UPF consumers was 86.4% (CI95 83.9 to 88.9).

**Figure 1.**
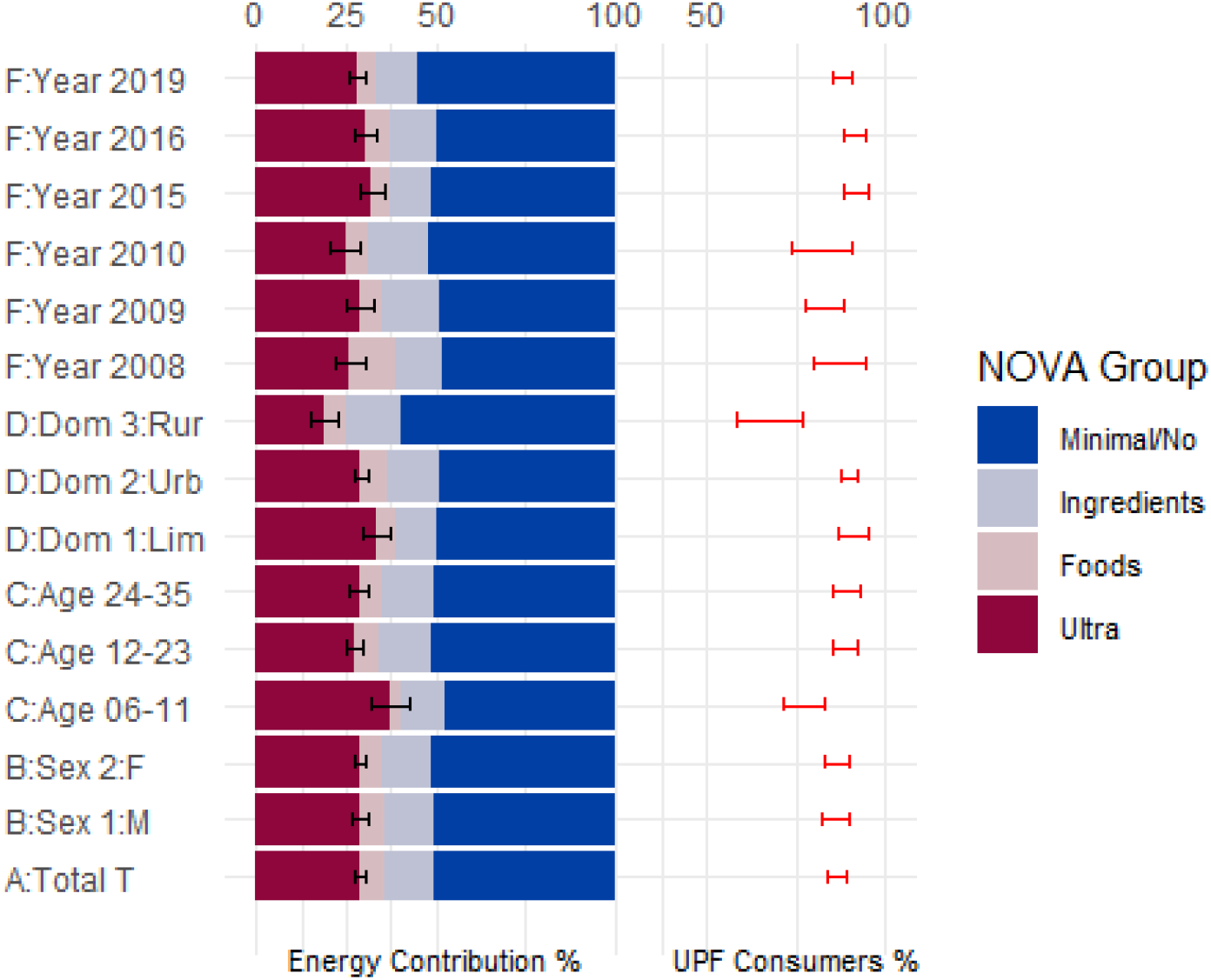
The left side of the figure shows stacked bars representing the total amount of energy (as % contributions) consumed for the total population and several covariate categories (vertical axis). The stacked bars show the weighed estimate for the energy contribution provided by the NOVA categories. For the UPF (NOVA 4) category 95% confidence bars are shown. The right side of the figure shows the percentage of children who did consume UPF. The horizontal scales for contributions and consumer percents are at the top. The total sample was n=2887 children.

The distribution of the UPF energy contribution by food group pointed to milk and derivatives as the main contributor with 18.6% (CI95 17.2 to 20.0) followed by cereals and derivatives with 4.5% (CI95 3.6 to 5.5). See figure in Supplement S7.

The square root transform of the UPF energy contribution has a clearly bimodal distribution with a distinct peak of non-consumers and a more or less symmetrical peak for UPF consumers (More detail in Supplement S2).

Table 2 shows the two models, normal for the mean transformed UPF energy contribution among UPF consumers only and binomial for the percentage of UPF consumers. Figure 2 presents a visualization of the models as a series of bi-variable slices (since no interactions have been detected), together with weighted estimations of the actual data. In the normal model, the mean UPF energy contribution non linearly decreases with age, increases with height-for-age, and gets a bit smaller in the Austral Summer and larger in Autumm. In the binomial model the percentage of UPF consumers clearly increases with age and height-for-age, slightly increases with survey year and decreases with poverty.

**Table 2.**

| Model | SlopeU | SlopeA | SlopeF | StdError | pValue | Description |
| --- | --- | --- | --- | --- | --- | --- |
| binomial |  | -107.779 | -84.958 | 50.030 | 0.090 | Intercept |
|  | 0.071 | 0.054 | 0.042 | 0.025 | 0.089 | Year |
|  | 0.010 | 0.020 |  |  |  | Day of Month |
|  |  | 1.066 | 1.038 | 0.206 | 0.000 | Age log |
|  | 0.426 | 0.275 | 0.294 | 0.087 | 0.001 | Height-for-Age z |
|  | -0.112 | -1.432 |  |  |  | Urban vs Lima |
|  | -1.555 | -1.649 |  |  |  | Rural vs Lima |
|  | -0.034 | -0.088 | -0.030 | 0.005 | 0.000 | District Poverty |
|  |  | 0.074 |  |  |  | Urban.Poverty |
|  |  | 0.058 |  |  |  | Rural.Poverty |
| normal |  | 211.511 | 10.694 | 1.768 | 0.000 | Intercept |
|  | -0.027 | -0.100 |  |  |  | Year |
|  | -0.000 | -0.670 |  |  |  | Day of Year |
|  | -0.020 | 0.128 | 0.134 | 0.043 | 0.002 | Age mo |
|  |  | -2.715 | -2.828 | 0.881 | 0.001 | Age log |
|  | 0.733 | 1.104 | 0.590 | 0.229 | 0.010 | Apr-Jun |
|  | 0.360 | 1.290 | 0.310 | 0.198 | 0.120 | Jul-Sep |
|  | 0.343 | 1.581 | 0.259 | 0.209 | 0.215 | Oct-Nov |
|  |  | -0.028 | -0.026 | 0.007 | 0.000 | HFAZ square |
|  | 0.320 | 0.295 | 0.293 | 0.044 | 0.000 | Height-for-Age z |
|  |  | 0.000 |  |  |  | Year x Day |

Outcome: Normal mean square root of UPF energy contribution among consumers (n=2531); Binomial log odds of UPF consumers (n=2887). Slope: U: unadjusted model with single covariables only. A: adjusted model with all automatically selected covariables. F: final model with selected covariables. Std. errors and p values shown for final model only.

**Figure 2.**
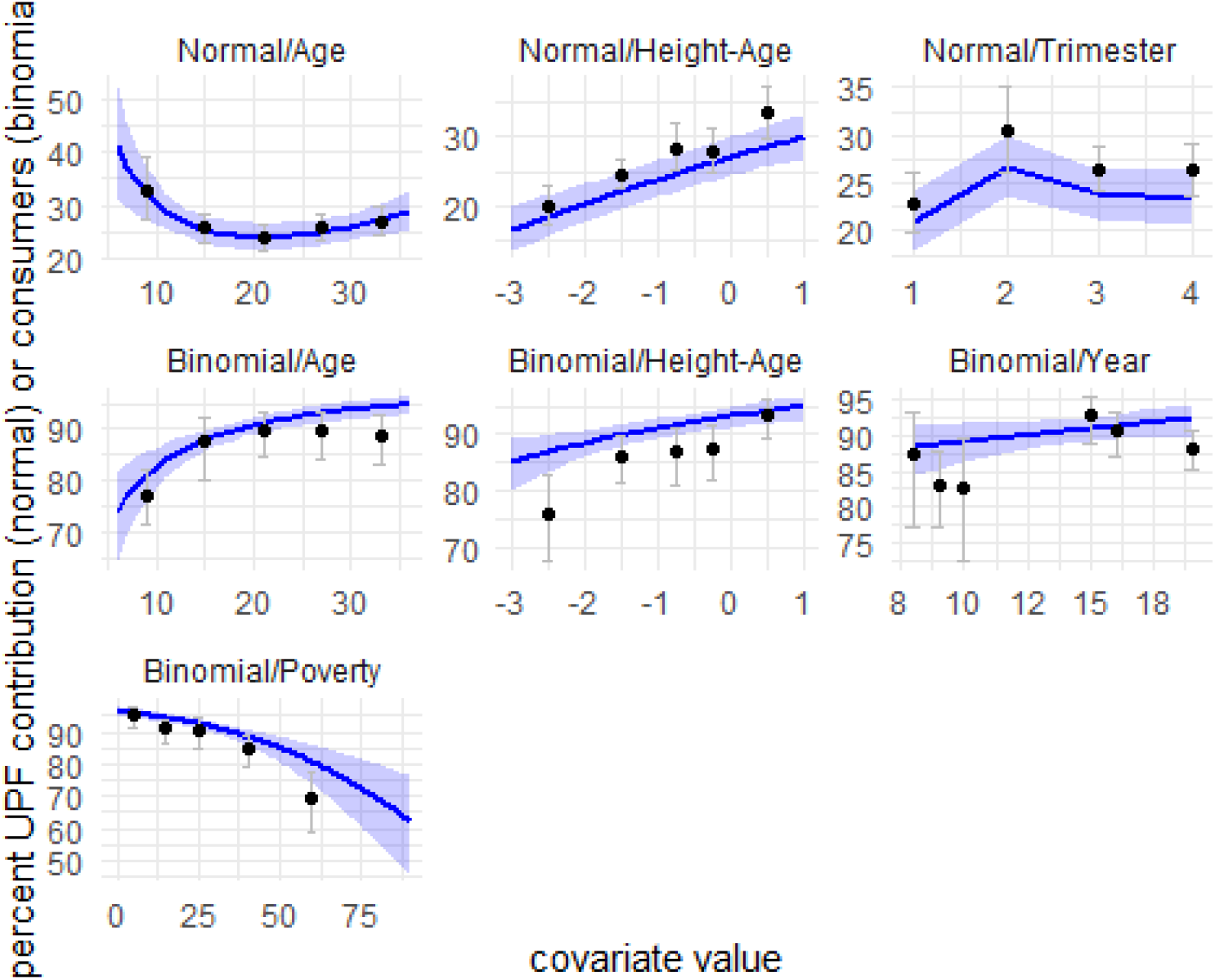
Each panel shows the scenario for a single covariate in each of the two models, Binomial (n=2887) lower four panels and Normal (n=2531) upper three panels. The outcome is in the vertical axis, percentage of UPF consumers for the binomial model, and mean UPF energy contribution among consumers for the normal model. The horizontal axis shows the covariate. The solid blue lines show the model prediction for an ‘average’ child (21 months age, -1 z HA score in the 3rd trimester of 2015, except for the varying covariate in each panel). The light blue shadows are the 95% confidence band for the model prediction. The black circles are the weighed estimate from the actual sample (therefore with the actual mix of covariates) and the gray bars are the corresponding weighed 95% confidence interval. No interaction terms were selected in the final models, so none is depicted in this figure.

## Discussion

These findings, from two surveys by the Peruvian Instituto Nacional de Salud, are the first estimation of UPF intake for children in Peru. Such intake is measured with two non-redundant indicators: the mean UPF energy contribution in consumers (30-33%) and the percentage of UPF consumers (84-89%). Our analysis also finds that the mean UPF energy contribution among consumers decreases, with age and height-for-age, and shows some seasonal variation (a bit smaller in the Austral Summer and larger in Autumm) while the percentage of UPF consumers increases with age, decreases with poverty and height-for-age, and shows a slight increase over survey years.

When comparing the point mean UPF energy contribution found here, 30% among 6-35m children with other countries^6^ for the nearest age group, 2-5y, we see Peru at 30 below Mexico 38, Chile 44, Australia 47, USA 58, UK 61, and above Colombia 18.

In other Latin American countries, with data of individuals over 2 years old, there is evidence that the UPF energy contribution decreases with age^7,9,10^ while our modeling finds a decrease between 6 and 23 months old (the age period where the percent of UPF consumers, while growing, is still below 90%), and seems to start an increase afterwards. A possible explanation could be the introduction of foods intended for small children, such as dairies (particularly evaporated milk) and cereals. The energy contribution from these foods would reduce afterwards while the household diet gets introduced. Our sample does not finely model the third year of life, while the literature does not show data of the first two years of life.

There is also some evidence^9,10^ for socioeconomic differences, which may be related to UPF pricing. Although plausible, we have not found literature on time trends, seasonal variation, or height-for-age differences of UPF energy contribution in children. The seasonality could be related to UPF availability, migration, climate, income or expenses, hypotheses that deserve further study. The height-for-age differences could be related to early undernutrition, a consequence of economic deprivations limiting access to UPF.

In this age group we find that most of the UPF intake comes from milk and cereal products. Evaporated milk is the most common UPF, consumed by 58% (CI95 55 to 61) of children. Fresh milk is consumed only by 7% (CI95 6 to 9). Evaporated milk provides several macro and micronutrients, besides having sodium, saturated fats and food additives and perhaps promotes intake of added sugars ^21,22^. Evaporated milk does not require refrigeration until the can is opened. Evaporated milk has also been an important industry in Peru since about 1940^23^ . Thus, the issue of advisability of evaporated milk needs further research.

Food labelling norms in Peru, requiring octagon risk symbols in packaging, were enacted in 2018 ^24^. It will be important to watch the UPF trends in the following years.

Among the limitations of our study we have (1) representation of rural areas (although formally included and properly weighted, we have the impression that field difficulties may have generated poorly documented losses); (2) missing recall data (being a long procedure requiring appointments for the randomized days); and (3) reproducibility of Nova classification, especially for UPF. Some of these limitations may produce overestimates and some underestimates.

Among the strengths of our study we have (1) it covers all the regional diversity; (2) it achieves a sizable sample size, as reflected in the CI95; (3) it applies a fairly standardized R24h; (4) it applies the R24h on random days; and (5) it applies statistical adjustments for the bimodality (zero inflation), asymmetry, confounding and complex sample. This study could be a baseline for UPF intake of children in Peru before the implementation of front package labelling.

We conclude that, among 6-35 months-old children in Peru between 2008 and 2019, ultra-processed (Nova 4) foods (1) contribute 30% of energy among UPF consumers, a mean that decreases with age, increases with height-for-age, and shows seasonal variation; (2) are consumed by 84 to 89% of children, a proportion that increases with age, height-for-age, year and decreases with poverty; and (3) are provided mostly by milk or milk derivatives and cereals or cereal derivatives.

We recommend that, given the increasing evidence against UPF, the time trend should be watched also in this age group. The advisability of discouraging evaporated milk and cereals should be reviewed. A deeper analysis of the UPF pattern in 6-59 months old should be carried out, that being a very dynamic period in life.

## Data Availability

The original data is available at government sites as specified in the supplement.
The anonymized analytic dataset and the code is available as open data specified in the supplement.

https://www.datosabiertos.gob.pe/dataset/monitoreo-nacional-de-indicadores-nutricionales-monin-cenan-2007-2010

https://www.datosabiertos.gob.pe/dataset/monitoreo-nacional-de-indicadores-nutricionales-monin-cenan-2007-2010

https://www.datosabiertos.gob.pe/dataset/encuesta-vianev-ni%C3%B1os-2016-estado-nutricional-consumo-de-lm-y-consumo-de-alimentos-ins-cenan

https://www.datosabiertos.gob.pe/dataset/encuesta-vianev-2019-estado-nutricional-y-consumo-de-alimentos-del-ni%C3%B1o-menor-de-5-a%C3%B1os-de

## Acknowledgements

The MONIN and VIANEV surveys were funded by the Peruvian Instituto Nacional de Salud and received support from the Peruvian Instituto Nacional de Estadística e Informática. MONIN also had partial support from the International Development Bank, the World Bank and the US Agency for International Development. VIANEV also had partial assistance from the Peruvian Ministerio de Desarrollo e Inclusión Social and the Ministerio de Educación. Both surveys were carried out by teams dedicated to the careful application of food intake surveys. We thank them all.

