## supplemental file for "Energy contribution from ultra-processed foods in Peruvian children: Frequency, Bimodality and Determinants"

Version 2024-May-23.

By:

Marianella Miranda-Cuadros,

Miguel Campos-Sánchez

Gustavo Cediel

María Laura da Costa Louzada

Joaquín Alejandro Marrón-Ponce

#### Section S1 NOVA Classification

Quoting from the recent FAO guide (Monteiro et al 2019a):

> The NOVA classification system groups all foods according to the nature, extent and purposes of the industrial processes they undergo. These involve physical, biological and chemical techniques used after foods are separated from nature, and before they are consumed or else made into dishes and meals.

From that publication´s Table 1 in the Spanish version the adaptation in the following Table S101 has been made, incorporating clarifications from other sources (Monteiro et al 2019a, Monteiro et al 2019b, FAO 2021, PAHO 2019). From this guideline, the sequence of programmed rules was prepared, as mentioned in the main article. Table S102 presents a verbal summary of that sequence of rules. Both tables. S101 and S102 are presented here in Spanish, as they were originally produced.

##### Table S101 Definitions of NOVA Classification

| |
|---|
| Group 1: Unprocessed or minimally processed foods. |
| Unprocessed: edible parts of plants (seeds, fruits, leaves, stems, roots) or of animals (muscle, offals, eggs, milk), and also fungi, algae and water, after separation from nature.<br>Minimally processed: unprocessed foods altered by industrial processes such as removal of inedible or unwanted parts, drying, crushing, grinding, fractioning, filtering, roasting, boiling, pasteurisation, refrigeration, freezing, placing in containers, vacuum packaging, or non-alcoholic fermentation. The main aim of these processes is to extend the life of grains (cereals), legumes, vegetables, fruits, nuts, milk, meat and other foods, enabling their storage for longer use, and, often, to make their preparation easier or more diverse.<br>In none of these processes salt, sugar, oils or fats or sweeteners are added to the original foods.<br>Foods with vitamins and minerals added generally to replace nutrients lost during processing, such as wheat or corn flour fortified with iron and folic acid.<br>Group 1 foods do not usually contain additives to preserve their original properties or may have additives that prolong product duration, protect original properties or prevent proliferation of microorganisms.<br>The group includes herbs and spices used in culinary preparations, such as thyme, oregano, mint, pepper, cloves and cinnamon, whole or powdered, fresh or dried. |
| Group 2: Processed culinary ingredients. |
| Substances obtained directly from Group 1 foods or from nature by industrial processes such as pressing, refining, trituration, grinding extracting or mining and atomization drying. Normally they are not eaten alone, but are used as ingredients in the preparation, seasoning and cooking of group 1 foods, so that preparations, soups, breads, salads, drinks, desserts and other culinary preparations have a pleasant taste and become diverse, nutritive and satisfying.<br>Examples are mined or seawater salt, sugar and molasses from sugar cane or beet, honey extracted from combs and syrup from maple trees, vegetable oils pressed from olives or seeds, fat obtained from milk (butter) or pork (butter), starches extracted from corn and other plants (cornstarch), products having two elements from Group 2, such as salted butted, products consisting of two group 2 items, such as salted butter, and group 2 items with added vitamins or minerals, such as iodised salt, and vinegar obtained by acetic fermentation of wine and other alcoholic beverages. Elements of Group 2 may contain additives to preserve the original properties. |

| Group 3: Processed foods. |
|---|
| Relatively simple products made by adding sugar oil, salt, or other group 2 ingredients to group 1 foods. Most processed foods contain two or three ingredients. Processes include preservation methods such as canning and bottling, and, in the case of breads and cheeses, using non-alcoholic fermentation.<br>Processes and ingredients here aim to increase the durability of group 1 foods, modifying or enhancing their sensory qualities.<br>Some examples are canned or bottled vegetables, fruits and legumes in brine; salted or sugared nuts and seeds; salted, cured, or smoked meats (ham, bacon, pastrami); canned fish with oil, salt or smoked; fruits in syrup; and most freshly made unpackaged breads and cheeses with salt.<br>Processed foods generally preserve their basic identity and most original constituents. These products may contain additives that prolong product duration, protect original properties or prevent proliferation of microorganisms. Some examples are fruit as syrup with added antioxidants and dried salted meats with added preservatives. |
| Group 4: Ultra-processed foods. |
| Industrial formulations, generally having five or more ingredients. Besides salt, sugar, oil and fat, foods not usually employed in culinary preparations are included, such as sugar varieties (fructuose, high fructose corn syrup, fruit juice concentrates, inverted sugar, maltodextrin, dextrose, lactose), modified oils (hydrogenated or interesterified oils) and protein sources (protein hydrolisates, soy protein isolate, gluten, casein, serum protein and mechanically processed meat) and additives used to emulate the sensorial properties of unprocessed of minimally processed foods and culinary preparations or to hide undesirable properties of the final product such as colours, flavours, flavour enhancers, sweeteners (aspartame, cyclamate or stevia derivatives), emulsifiers (thickeners), emulsifying salts, humectants, sequestrants, firming agents, bulking, anti-foaming agents, anti-caking agents, gelling agents, coating and glazing agents. They are commonly referred to as a class, like flavouring, natural flavors or artificial flavor. Or their names are followed by their class, like "monosodium glutamate (flavor enhancer)" or "caramel color" or "soy lecitin as emulsifier". In any case, the UN Codex Alimentarius provides an updated list of additives with their functional classes as well as an online search service.<br>The main purpose of ultra-processed foods is to create ready to eat, ready to drink or ready to heat products, capable of substituting unprocessed or minimally processed foods as well as freshly prepared dishes.<br>Some examples of ultra-processed foods are carbonated soft drinks; sweet, salty or savoury packaged snacks; ice cream, chocolate, candies (confectionery); mass-produced packaged breads and buns; margarines and other spreads; processed cheese, cookies (biscuits), pastries, cakes, and cake mixes; breakfast 'cereals', 'cereal' and 'energy' bars; 'energy' drinks; milk drinks, 'fruit' yoghurts and 'fruit' drinks; 'cocoa' drinks; 'instant' sauces; meat or chicken extracts; infant formulas, follow-on milks, other baby products; 'health' and 'slimming' products such as meal replacement shakes and powders. Many ready to heat products including pre-prepared pies and pasta and pizza dishes; poultry and fish 'nuggets' and 'sticks', sausages, burgers, hot dogs, and other reconstituted meat products, and powdered and packaged 'instant' soups, noodles and desserts. Their ingredients and formulations during pre-elaboration make them ultra-processed.<br>Products prepared exclusively with Group 1 or Group 3 foods which also contain cosmetic additives or sensorial enhancers, like natural yogurt with artificial sweeteners and breads with emulsifiers are included in Group 4. |

#### Table S102 NOVA Categories for Foods and Drinks

| | Food Group | Description | Nova Group |
|---|---|---|---|
| A | Cereals and derivatives | | |
| | | grains or seeds, whole, broken or peeled: quinoa, wheat, barley, corn, rice, etc | 1 |
| | | grains or seeds presented in flour, semolina, polenta | 1 |
| | | grains or seeds presented as flakes | 1 |
| | | noodles (except instantaneous) | 1 |
| | | cereal flakes, fortified and precooked | 1 |
| | | cornstarch | 2 |
| | | cereal flake, fortified and precooked, with sugar | 3 |
| | | "bread" or similar and pastries (without brand) freshly baked | 3 |
| | | "bread" or similar and pastries (branded) | 4 |
| | | cereal products: Cerevita, Kiwigen | 4 |
| | | pan de "molde" | 4 |
| | | cakes, from store | 4 |
| | | Cookies | 4 |
| | | “panetón” | 4 |
| | | toasts | 4 |
| | | energy bars, with cereals | 4 |
| | | “barquillos” | 4 |
| | | cereal flake, fortified and precooked, with sugar and additives | 4 |
| | | corn flakes | 4 |
| | | packed infant cereals (Nestum, Cerelac, etc) | 4 |
| B | Vegetables and derivatives | | |
| | | vegetables, natural or cooked | 1 |
| | | pickled or brine (with preservatives) | 3 |
| | | tomato sauce (without additive or with preservative like citric acid | 3 |
| | | tomato sauce, concentrated, with meat or not (with other additives), ketchup | 4 |
| | | pickled or brine (with preservatives and other additives) | 4 |
| C | Fruits and derivatives | | |
| | | fruits, natural or cooked | 1 |
| | | fruit (100%) juice or “agua”, or pasteurized, without sugar | 1 |
| | | fruit juice, pulp | 1 |
| | | fruit juice, sweetened (with preservative like citric acid) | 3 |
| | | fruit pulp nectar, sweetened | 3 |
| | | nectar (with additive, preservative like citric acid) | 3 |
| | | nectar (with additive like citric acid, stevia or other) | 4 |
| | | fruits in syrup, candied fruit | 4 |

| | | | |
|---|---|---|---|
| D | Fats, oils and oil products | | |
| | | peanuts and other oleaginous | 1 |
| | | cocoa | 1 |
| | | animal fat | 1 |
| | | Butter (including “manteca”) | 2 |
| | | oil | 2 |
| | | peanut butter | 3 |
| | | peanut butter (with additives) | 4 |
| | | chocolate | 4 |
| | | products like Milo, Nescao or similar | 4 |
| | | margarine | 4 |
| | | candied peanut | 4 |
| E | Fish and seafood | | |
| | | fish or seafood, natural or cooked | 1 |
| | | sardine, tuna or other, canned/”conserva”, with oil, salt or tomato sauce | 3 |
| | | fish or seafood, dry or salted | 3 |
| | | fish or seafood, dehydrated or cured | 3 |
| F | Meats and derivatives | | |
| | | meat, any animal, raw or cooked | 1 |
| | | meat, half-dried, dried, smoked, fried or salted | 3 |
| | | charqui/chalona/charque | 3 |
| | | bacon, “tocino” or “Cecina” | 3 |
| | | meat or derivatives, canned (with salt or oil) | 3 |
| | | hamburguers, hot dogs, sausages, ham | 4 |
| | | meats, breaded like “nuggets” | 4 |
| G | Milk and derivatives | | |
| | | milk, animal, whole, natural, or fresh | 1 |
| | | curdled milk, milk serum | 1 |
| | | milk, "pasteurized" or "ultrapasteurized", liquid | 1 |
| | | natural yogurt (milk and cultures, like Danlac brand) | 1 |
| | | cheeses (with salt) | 3 |
| | | “quesillo” | 3 |
| | | yogurt and dairy drinks, flavoured and sweeteneds, culture milk | 4 |
| | | milk, "saborizada" | 4 |
| | | milk cream | 4 |
| | | condensed milk | 4 |
| | | melted cheese | 4 |
| | | milk "evaporated" (whole, skim, semiskim, light, lactose-free, UHT) | 4 |
| | | cream cheese crema, melted | 4 |
| | | natural yogurt (with additive like pectin) | 4 |
| | | milk, canned “chocolatada” (Gloria, Laive…) | 4 |
| | | “manjarblanco” | 4 |
| | | powder milk (with additives like Gloria, Pura Vida, etc) | 4 |
| | | dairy mix or product, milk substitutes | 4 |

| | | | |
|---|---|---|---|
| H | Alcoholic and Nonalcoholic Beverages | | |
| | | without sugar | 1 |
| | | beer | 3 |
| | | wine | 3 |
| | | nectar (pulp, juice, water and sugar) | 3 |
| | | nectar (with additive: citric acid) | 3 |
| | | nectar (with additive like citric acid, stevia, others) | 4 |
| | | rum, cognac, similar | 4 |
| | | kola (cola), soda | 4 |
| | | milk, canned “chocolatada” (Gloria, Laive…) | 4 |
| | | oral rehydration solution | 4 |
| | | packed, canned or bottled soy drink | 4 |
| J | Eggs and derivatives | | |
| | | eggs, fresh or any degree of cooking | 1 |
| | | eggs, dehydrated, dried | 1 |
| K | Sugar products | | |
| | | sugar cane | 1 |
| | | honey, “chancaca” | 2 |
| | | sugar | 2 |
| | | syrup, jelly (“jalea”) | 2 |
| | | marmalade | 4 |
| | | jelly (“gelatina”) | 4 |
| | | cotton candy | 4 |
| | | powdered sugar | 4 |
| L | Miscellaneous | | |
| | | annatto (“achiote”), algae, saffron, “yuyo” algae, cocoa, cumin, carob powder, mushrooms (“champiñón”, “seta”), laurel (bay leaf), pepper, anise, clover, cynammon, nuts, kion, cushuro, powdered barley | 1 |
| | | tea (leaves) , coffee (grain or grinded) | 1 |
| | | salt | 2 |
| | | yeast | 2 |
| | | vinager (non syinthetic) | 2 |
| | | synthetic vinager (ethanol) | 4 |
| | | “sillao”/soy sauce | 4 |
| | | "in syrup" | 4 |
| | | meat/chicken/hen broth | 4 |
| | | ketchup, jelly | 4 |
| | | mayonnaise | 4 |
| | | packed sauces | 4 |
| | | powdered coffee | 4 |
| | | cereal bar | 4 |
| | | Ice cream | 4 |

| | | | |
|---|---|---|---|
| Q | Infant Foods | | |
| | | porridge from Social Programs | 1 |
| | | porridge or strained “colado” (Heinz and similar) | 3 |
| | | breast milk substitutes | 4 |
| | | cereales en lata (nestum, cerelac, etc) | 4 |
| T | Legumes and derivatives | | |
| | | all beans or legumes, including their flour | 1 |
| | | canned soaked bean (without aditivo) | 3 |
| | | canned “menestra” drinks or "milks" | 4 |
| U | Tubers, roots and derivatives | | |
| | | natural or cooked | 1 |
| | | flour or powder | 1 |
| | | dehydrated | 1 |
| | | mashed dehydrated | 4 |
| | | flake for mash | 4 |
| V | Andean tubers | | |
| | | natural or cooked | 1 |

#### Section S2 Distribution of the Outcome Variable

Initially, the outcome variable was the energy intake fraction provided by UPF, that is, the total energy provided by UPF divided by the total energy intake, both per 24h.

##### Figure G002 Ultra processed Food Intake (Weighted)

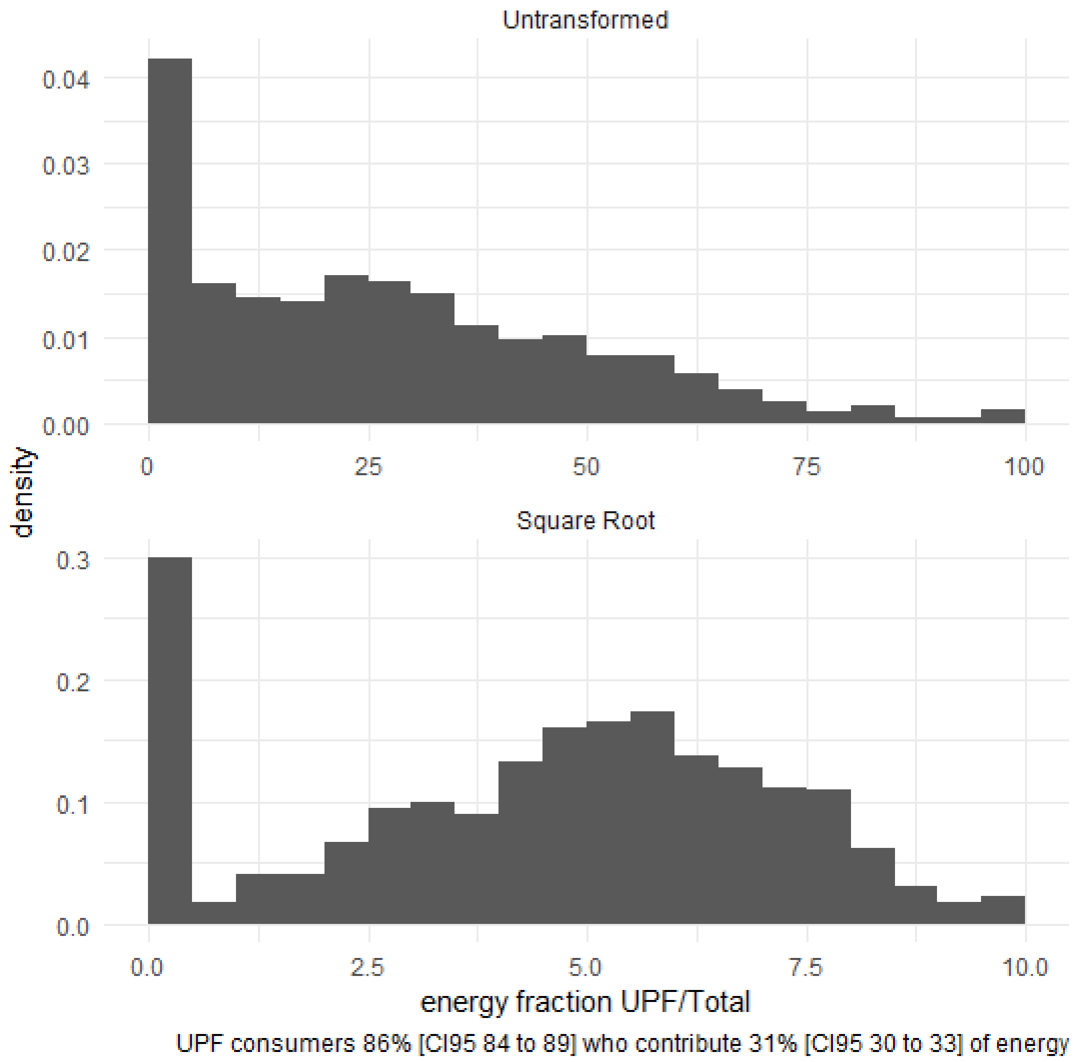

The upper panel shows the frequency distribution of the untransformed energy fraction among the 2887 children, adjusted for the complex sampling design. The horizontal axis has the energy fraction, expressed as percentage. The vertical axis has the frequency, expressed as the probability density. The lower panel shows the frequency distribution of the square root of that energy fraction, adjusted for the complex sampling design.

The transformed variable is clearly bimodal. The left peak has 14% [CI95 11 to 16] children who were not UPF consumers. The right peak has the remaining 86% [CI95 84 to 89] children who were UPF consumers.

#### Figure S002 Ultra processed Food Intake Transformations (Unweighted)

To arrive at the square root transformation, Box-Cox (in four variants) and Tukey transformations were fitted to the unweighted data for consumers only, obtaining the following lambda and gamma coefficients for the best fit in each transformation.

| Method | Package | Function | Option | Lambda | Gam | Min | Max |
|---|---|---|---|---|---|---|---|
| Box & Cox | car | powerTransform | bcPower | 0.490 | NA | -2.02 | 0 |
| Yeo & Johnson | car | powerTransform | yjPower | -1.35 | NA | 0.0000495 | 0.451 |
| Hawkins & Weisberg | car | powerTransform | bcnPower | 0.262 | 0.1 | -2.08 | 0.00249 |
| Tukey | rcompanion | transformTukey | <NA> | 0.625 | NA | 0.00204 | 1 |
| Box & Cox | MASS | boxcox | <NA> | 0.45 | NA | -2.20 | 0 |

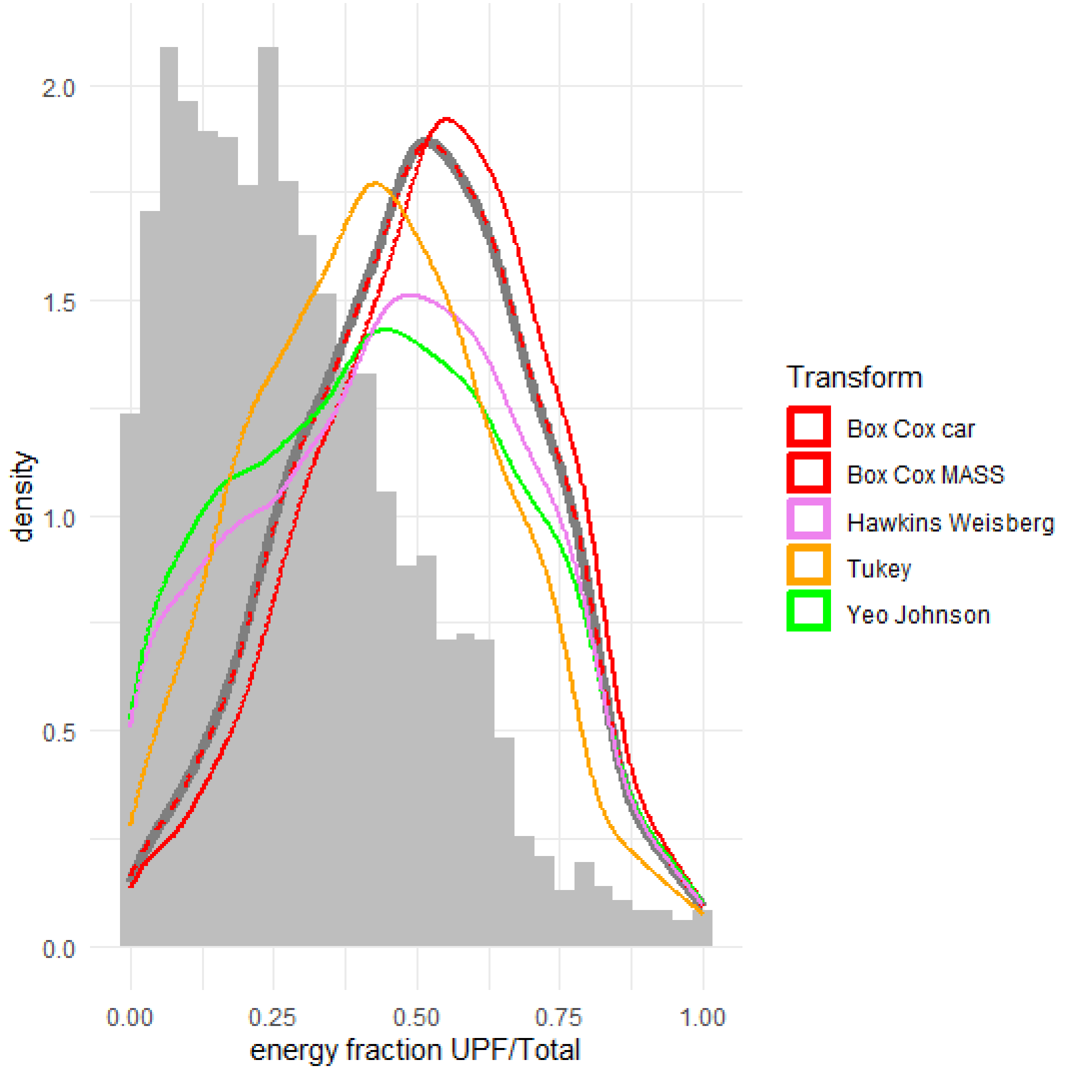

The density distributions of the original and transformed (rescaled to the range [0,1]) variables are shown. The square root transformation has been added, and finally chosen, because the Box -Cox lambda estimates are close to 0.5 and it is a simple transform.

### Section S3 Data Processing Techniques

#### Input Files

The input files for the processing are listed below, together with their MD5 signatures:

```
   PART  FNAM                                                   BYT MD5                              FEC
   <chr> <chr>                                                <dbl> <chr>                            <chr>
 1 LIBS  NuTabs.rda                                          594646 66404cdac1771bb9d3bda6a42cf10393 2023-02-01 10:18:26
 2 LIBS  GCTabs.rda                                          320674 dad57e6b3fbb948d0a318a51a2e6d1d9 2021-08-13 13:46:11
 3 LIBS  PETabs.rda                                          722325 a707649f46cf49ace28ed05ee4983774 2023-08-13 22:39:52
 4 LIBS  TUBIGC.csv                                          211997 163f17d6c7b786580b490008b3d085e0 2023-09-08 12:17:44
 5 VN15  BD.01.Caratula-Caract.Viv.sav                       648123 c91e092330d885e5658623b815e50c59 2018-01-30 08:34:52
 6 VN15  BD.03.Salud.y.Nutricion.sav                         160411 e2e843c8cea13643b05890c31fe9ca88 2018-01-30 08:37:01
 7 VN15  BD.02.Caract-Miemb.Hogar.sav                        278996 9ac40fc1bcf471d2b22393d507d9aab2 2018-01-30 08:36:18
 8 VN15  BD.09.Consumo.Ind-Niño-Hogar.sav                   7500562 a15568088efc62522999cdd983493480 2018-01-30 08:42:38
 9 VN15  BD.05.Consumo.Ind-Niño-Cuna.Mas.sav                  90981 7f5058cff8319565dd8562f0575f22c8 2018-01-30 08:39:31
10 VN15  BD.10.Preparacion.Ind-Niño-Hogar.sav               6596076 3a5003a1b93f6656828587e3b44efc6f 2018-01-30 08:43:02
11 VN15  BD.06.Prepar-Cuna.Mas.sav                           187642 ed622a5375cd7330c6b25f29e5b13234 2018-01-30 08:42:04
12 VN16  CAP001.sav                                          788455 54c33b08eb37fa1a7023143629daf773 2017-03-16 22:39:24
13 VN16  CAP200.sav                                          567101 c589c1e4a626a87730f249f195cb2e2c 2017-03-16 22:49:59
14 VN16  CAP400.sav                                          321462 f27074330d0eb1eccc1dcc1530b622bf 2017-04-13 15:44:30
15 VN16  CAP618.sav                                         4284755 7466a6285952517cd363129cf582dd02 2017-04-13 22:30:23
16 VN16  CAP600.sav                                           34149 c936dfca59bcfdad83bdf02540dfc49a 2017-03-16 23:06:50
17 VN16  CAP623.sav                                         4548241 a4795d9125bab3e5d5483a0419f9535d 2017-04-13 23:25:05
18 VN16  CAP606.sav                                          100013 e39ed65539793efd3bf933945d368856 2017-03-17 14:38:32
19 VN19  NIÑOS 2019 INDI CONSU HAB.sav                       1613211 cb1cde5f20815af218f13198f1f645d2 2023-09-19 20:56:17
20 VN19  CAP 621 CONSUM INDIV HOGAR1.xlsx                   5252152 775a797e198ae00161d07683f316a270 2023-09-19 20:56:09
21 VN19  CAP 600 CONSUM INDIV CUNA MAS.xlsx                   42779 2ee4bfb3b3fd13f1ffbabf1eed9b6ff9 2023-09-19 20:56:09
22 VN19  CAP 627 PREP ELAB HOGAR_ CONSISTENCIA MINIMA.xlsx 11568742 0d6a8f2710399e30eafe38b2cbeda354 2023-09-19 20:56:09
23 VN19  CAP 607 PREP ELAB CUNA MAS.xlsx                     141326 de88ad61fb6e2e2c373fb57cacc6ef1b 2023-09-19 20:56:09
24 VN19  L2022123019v.rds                                      5148 6abc489064b0d65ec960c006a0cd3050 2023-09-19 20:59:17
25 MON3  V12.MDB                                           31039488 667937e87ebe8a8aee0e3c8376875334 2012-12-06 12:33:24
26 MON3  WT.sav                                             2066651 441c24dd757bcd0e7cd8d827635dff68 2014-04-13 21:45:56
27 MON3  P10.sav                                            1710283 b4385287c6c29fc8080e0f9efbbc86f8 2010-12-21 14:20:12
28 REFS  03 TABLA DE ALIMENTOS2016.xlsx                      649086 7a672c95aa1f21670ecb56b97a8ffda5 2021-04-30 13:35:56
29 REFS  TABLA DE ALIMENTOS.sav                              469462 2a6aa4dd8dbc20d3d8437e6216a25cc0 2021-07-06 18:34:54
30 REFS  TABLA ALIM INDUSTR.sav                              258736 cacd57ff68d093a2eb2ca1ad8e83bb5d 2021-07-06 18:36:38
31 REFS  L2023020109_sintaxis en excel11g.xlsx                33776 9672c464db5bd364791c9006a667bd60 2023-03-19 09:34:10
32 REFS  L2023112217_NOVA  preparaciones -221123.xlsx         56051 05194b31a8510e71b32a0162b820cecc 2023-11-23 17:24:31
```

The PART=LIBS files are publicly available sets of several reference tables and R functions built by us for several data analysis tasks, apart from this article (Campos 2021).

The NUTABS file contains consolidated food composition tables for Peru (INS/CENAN 2017, PRISMA 2003), and household measure equivalence tables (Miranda et al 1996) as well as collected food equivalence tables used by the MONIN III survey (INS/CENAN 2023).

The NUTABS file also contains reference tables and functions for computing the recommended dietary energy requirements (FAO/WHO 2004 and IOM 2005).

The GCTABS file contains consolidated anthropometry tables and functions for the WHO Reference Data (WHO 2006).

The PETABS file contains several consolidated tables for Peru, from which we have used in this article the official population projections (INEI 2009) and the poverty maps (INEI 2010, 2015 2020).

The PART=VN15 files are consolidated data for the VIANEV 2015 survey (INS/CENAN 2023). The PART=VN16 files are consolidated data for the VIANEV 2016 survey (INS/CENAN 2023). The PART=VN19 files are consolidated data for the VIANEV 2019 survey (INS/CENAN 2023). L2022123019v is a patch file containing the district code for some households with missing data (but cluster identification number).

The PART=MON3 files are consolidated data for the MONIN survey which had food intake recalls between 2008 and 2010 (INS/CENAN 2023).

The PART=REFS files contain food composition data which were added to the consolidated NUTABS files from added foods in the VIANEV surveys and the rules for NOVA classification (a main rules file and a patch for initially unclassified foods, mainly drinks and prepared meals).

The MONIN and VIANEV data files for this article may have some discrepancies with the data files as officially published in the references quoted above (official repository publication of open data has only been available for INS/CENAN since the second half of 2023). Our impression is that those discrepancies are annoying but minor, and due to some disorder in the file housekeeping. Some details follow.

The VIANEV data had not been made public by the time this article was being prepared. Only some reports had been produced (INS/CENAN 2018, 2021, 2023) but data was publicly available only for 2015. In Peru such files can be obtained by any citizen through a Transparency of Information request, but they also can be obtained through internal official channels within INS/CENAN, which is the way we had access. The data we received had some differences in data structure between years, and there were some remaining inconsistencies which were removed during the Load step.

The MONIN surveys are three cycles, MONIN I 1995-2002, MONIN II 2004-2006 and MONIN III 2007-2010 (Campos et al 2011), only the last one has been used here. The reports and data were published initially in the CENAN web site under the denomination “Biblioteca Digital en Nutrición” (BDN) where it was available for several years. That site was taken down and replaced by links in the INS/CENAN/DEVAN web site, which has been also made inactive. The data has just recently been made available again in the repository (INS/2023). In each incarnation the data files have different structures because of different ways of compiling the information. For this article we have used one of the first versions, which was stored at the BDN, supplemented by a version of such file which was prepared for the MONIN report on food intake (Miranda et al 2012) and a research thesis (Miranda 2014).

Some of the input files listed here do have personal identification information which has been dropped in the ETL process.

#### Processing Workflow

In this section we provide a succinct description of the processing steps carried out to prepare the data before analysis. Such processing had two phases:

In the first phase, the original input files were read and consolidated files were produced. The steps during this phase were:

- Reading of VIANEV source files, with renaming and retyping of columns as necessary to have a common data structure for the three VIANEV rounds. The geographical coding for the districts was assigned here, on the basis of the region, province and district names (the first three administrative divisions of the country). No recoding or filtering were deemed necessary at this step.
- Consolidation of all VIANEV children files. Extremely low values for height (below 40 cm) and weight (below 1 kg) were recoded to missing. Age and WHO 2006 anthropometric indices were calculated. Exclusion of subjects who were outside the age range or did not have sampling weight information was carried out in this step.
- Patching of food codes for VIANEV 2015 and 2016 data, as detected by a cleaning checkout and decided upon by data inspection. All the patches are coded, and therefore registered, in the program. Most of these corrections are related to the type of presentation of the food (for instance dry versus fresh) and some clear misdoings to very different foods.
- Pooling of reference composition tables. From previous studies we had a pooled table, containing mainly several editions of the CENAN tables as well as the ANDREA table. To this base the codes for new foods registered during the VIANEV surveys were added. Mostly industrialized products for which composition was recorded from the food package labels or the manufacturer web sites. Food codes were reassigned in some cases where they overlap. The NOVA classification was applied here from the rules file.
- Consolidation of VIANEV food intake data into three intermediate files, for preparations, ingredients, and items (served or consumed). The weight as recorded in the VIANEV files has been used, without checking for household measure equivalences. Items with zero weight or lacking subject or R24 identification data were excluded. From the three intermediate files a consolidated file has been prepared containing both the items directly consumed (for instance a fruit), the ingredients directly recorded (for instance the items in a sandwich or a hot drink) or those whose weight was calculated from the household preparations and ingredients (for instance a soup, whose recipe is recorded for the family, and a certain amount has been served to the child). For meals taken at the Cuna Más social program, the composition is available. For some meals prepared outside the household (for instance from a restaurant), a reference composition is available.
- Calculation of nutrient intake for each child, essentially joining the consolidated food intake and food composition and classification tables and grouping them by child.
- Reading of MONIN source files. Since the MONIN survey had a separate, more detailed processing, most of the consolidation described for VIANEV was already done. MONIN did use the consolidated food composition table which was expanded with the new foods recorded during the MONIN survey. So, at this step only homolgation of column names and types was performed.

In the second phase, the consolidated files were used to prepare additional variables necessary for the analysis. The steps during this phase were:

- Consolidation of MONIN and VIANEV surveys.
- Computation of variables and terms for analysis and modeling.
- Joining with the district poverty maps.
- Exclusion of food with invalid weight and recordings of breast feeding intakes.
- Computation of the UPF energy intake variables (consumers and energy fraction).
- Computation of transformations of nutrient intakes divided by energy intake.
- Computation of FAO/WHO and USDA DRI energy requirements.
- Joining with yearly national population projections.
- Readjustment of sampling weights to the population projections.
- Computation of the Inclusion Condition (outcome, poverty and anthropometry).

The separation between both phases is just practical. The second phase is intended to be repeated more often during the analysis, as requirements for variables could arise.

#### Software R packages

The following R packages (and their pre-requisites) are specifically called by the load and analysis program:

Framework: tidyverse.

Parallel Processing: future, future.apply, furrr, parallel.

Data Management: dplyr, haven, labelled, readxl, DBI, odbc, openxlsx, readr, lubridate.

Complex Samples: survey, srvyr, svydiags.

Modeling: MASS, lme4, car, rcompanion, broom, DHARMa.

Table Production: gtsummary, flextable, gt.

Graphics Production: ggplot2, colorspace, GGally, cowplot, ggmosaic.

#### Data Availability

The management and analysis program R source code in a single file and the anonymized binary file containing data frames (LDAT) and estimates (LEST) are available under an open source license at https://github.com/vipermcs/pdata . Documentation for the frames is described at the end of the program and dictionaries are embedded (labelled) in the data frames (tibbles). The  current reference files NUTABS, GCTABS and PETABS are also available under and open data license at https://github.com/vipermcs/btools .

#### Section S4 Statistical Modeling of Risk Factors

The goal of the modeling process was to identify risk markers for the ultra processed food intake (UPF, i.e. NOVA 4 foods), as measured by a single 24-h recall (R24) interview.

Initially, the outcome variable was the energy intake fraction provided by UPF, that is, the total energy provided by UPF divided by the total energy intake, both per 24h.

Since the distribution of that variable was clearly asymmetric and bimodal, it was decided to build a set of two models for two outcomes:

- A discrete dichotomous variable, for all children, 1 for those children who ate UPF and 0 for those who did not.
- A continuous variable, for UP consumer children only, with the square root of the energy fraction (as a percentage).

The modeling technique is the generalized linear model (GLM) which is the multivariable logistic regression for the first outcome variable and the multivariable linear regression for the second variable. Both regressions were adjusted for the complex sample design.

The candidate risk factors, independent covariates, were represented as the following terms:

- EDM: age in months, with decimals (days between the interview date and the birth date, divided by 30.4375).
- BSEX: sex, dichotomous 1 male, 2 female.
- HFAZ: length-for-age z-score (WHO 2006 reference)
- WFHZ: weight-for-length z-score (WHO 2006 reference)
- IDDOM: geographical domain, categorical (Metropolitan Lima, Urban Rest and Rural)
- PPOB: district poverty prevalence de (INEI projections)
- DAEN: interview day of year, between 1 y 365
- AAEN: interview calendar year, between 2007 y 2019
- FTEN: interview trimester, categorical 1 to 4 (as a proxy for season)
- DSEN: interview day within the week, between 1 and 7
- DMEN: interview day within month, between 1 and 31
- SDEN: trigonometrical sin of 2.π.DAEN/365.25
- HFA2: HFAZ square
- WFH2: WFHZ square
- EDML: natural logarithm of EDM
- EDM:BSEX: interaction, EDML.BSEX product
- HFAZ:WFHZ: interaction, HFAZ.WFHZ product
- IDDOM:PPOB: interaction, IDDOM.PPOB product
- DAEN:AAEN: interaction, DAEN.AAEN product

The choice of square root transformation for the energy fraction outcome was based on a univariate Box-Cox adjustment whose exponent was close to 0.5. The transformed terms, such as squares, logarithms and trigonometrical functions, represent hypothesized non-linear relationships. The interaction terms represent hypothesized heterogeneities (synergies or antagonisms). This list of candidate terms has been discussed and agreed upon by the authors.

For each of the two models the following steps were carried out:

- Selection starting from the non-interaction candidate terms.
- Selection starting from the selected terms and the interaction terms.
- Verification of the selected terms.
- Diagnostic check of the final model.

Both selection steps were repeated AIC-based backward stepwise selections from 300 re-samples (using the original complex sample design) of the original data (bootstrap-like procedure to generate training samples). Those terms which were selected in more than 50% of the replications and had p-values less than 0.15 in more than 50% pf the replications were selected for the next step.

The verification step was the model fit to the original data (considered to be the testing sample), selecting for the final model those terms, single or interaction, which were statistically significant ($p<0.05$).

The diagnostic step was the examination of Cook's D scores for outliers, the examination of the variance inflation factors, both adjusted for the complex sample design, as well as the cumulated distribution of quantile residuals generated from a reweighed sample (a simple random sample of the same size as the original data obtained from the population expanded from the original data according to the sampling weights).

The GLM regression procedure uses classical techniques (McCullagh & Nelder 1989) with the necessary adjustments for the complex sample design (Lohr 2010, Lumley 2010). The selection procedure follows the advice to avoid overfitting (Harrell 2015) using a simplified implementation of bootstrap selection (Austin & Tu 2004, Rizopoulos 2023). The diagnostic procedure uses weighed estimations of D and VIF (Li & Valliant 2011, 2015, Liao 2010, Liao & Valliant 2012) as well as quantile residuals (Dunn & Smyth 1996, Austin & Tu 2004, Hartig 2022).

The software used was R 4.3.1. For GLM the svyglm function of the survey package was used. For the stepwise selection the stepAIC function from the MASS package was used. For diagnosis the svydiags and DHARMa packages were used. The implementation of the process took advantage of parallel R execution (future package) on a Windows and AMD platform.

### Section S5 Covariate Distributions and Associations

#### Figure G101 Distribution of the Sample over Time (Weighted)

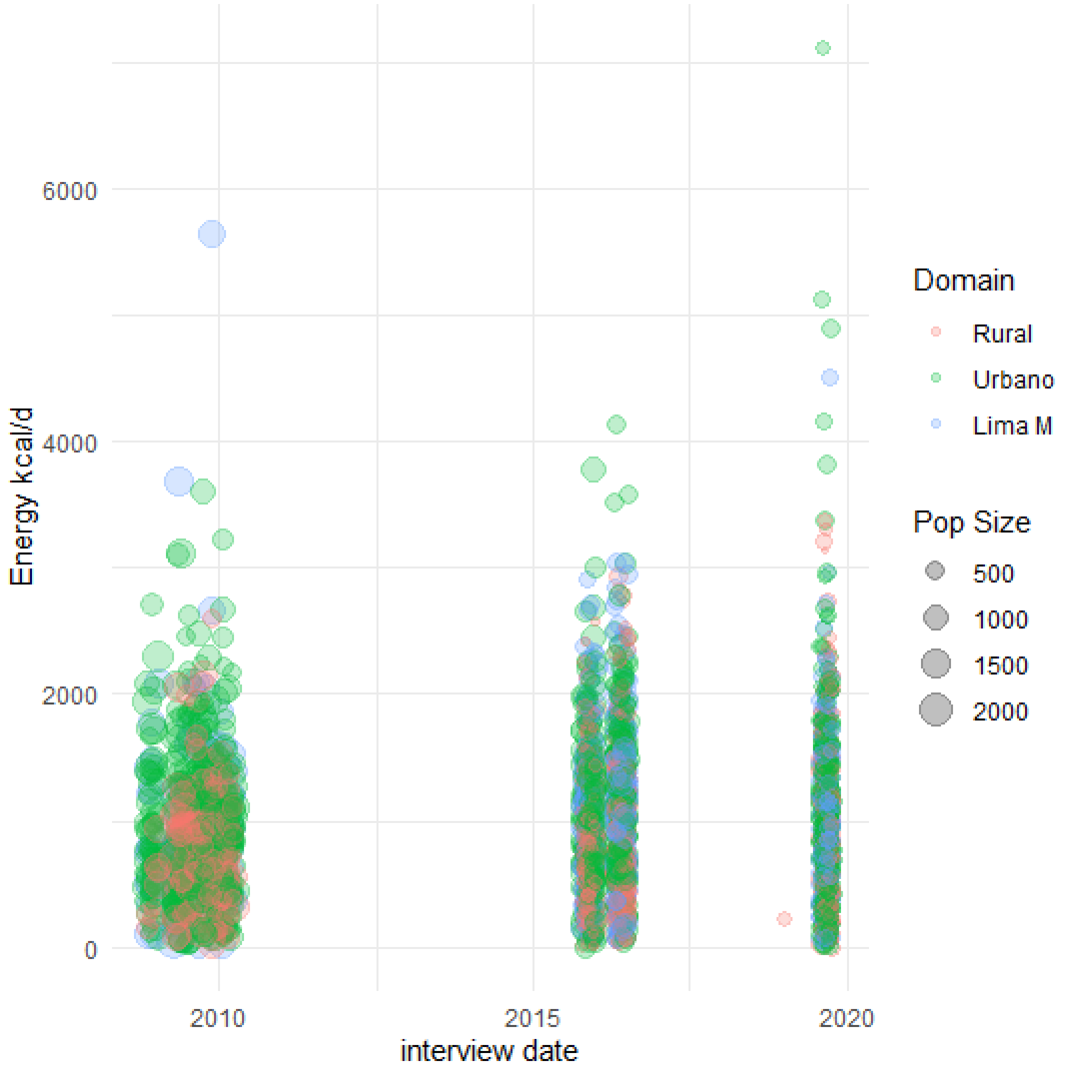

Each circle in the figure shows the energy intake value (vertical axis) and the date (horizontal value) for a sampled child. The circle diameter is proportional to the sampling weight. The colors correspond to the geographical domain. Notice that VIANEV surveys (2015, 2016 and 2018) do not cover a whole year and MONIN surveys (2008-2010) do not always cover whole years evenly.

#### Figure G201 Distributions of and Associations between Covariates (Unweighted)

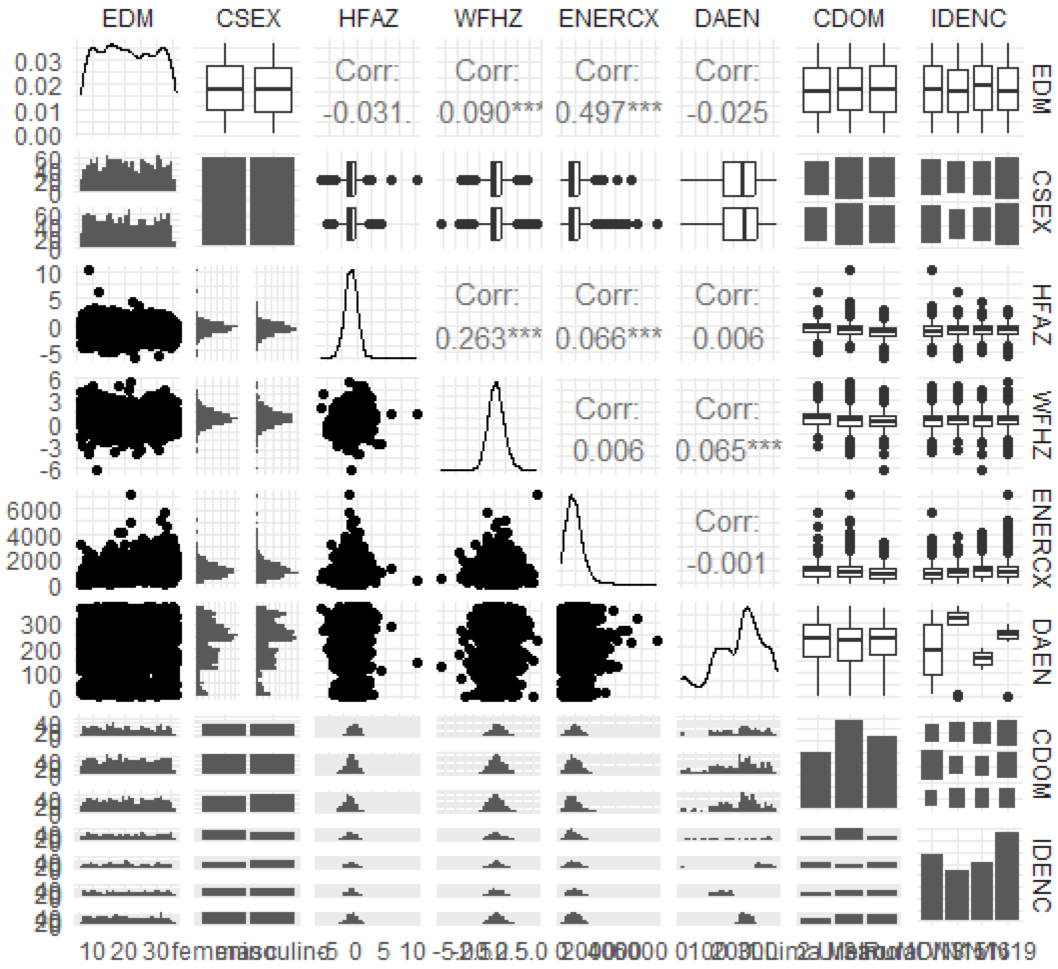

Each panel in the diagonal depicts the density distribution of a single covariate. Each panel apart from the diagonal depicts the correspondence between two covariates. Panels on the left-hand side below the diagonal show the same covariate pairs as those on the right-hand side above the diagonal, only transposed.

#### Section S6 Model Diagnostics

The transcript below shows the weighed diagnostic indices for both models (ISEQ=1 binomial and ISEQ=2 normal).

```
# A tibble: 2 × 12
   ISEQ NUOBS MCFAR2 COOKMD COOKQ1 COOKQ2  COOKP2  COOKP3 BETAPX LEVEP3 RESIP3   MCOVP0
  <int> <int>  <dbl>  <dbl>  <dbl>  <dbl>   <dbl>   <dbl>  <dbl>  <dbl>  <dbl>    <dbl>
1     1  2887 0.127  0.0553 0.0202  0.153 0.00277 0.00139 0.0187      0 0.0239 0
2     2  2531 0.0742 0.443  0.193   1.60  0.222   0.166   0.0269      0 0      0.000275
# A tibble: 13 × 11
    ISEQ TERM             TVAL  BETX    svy.vif   reg.vif       zeta        varrho zeta.x.varrho R.square FGT3
   <int> <chr>           <dbl> <int>      <dbl>     <dbl>      <dbl>         <dbl>         <dbl>    <dbl> <lgl>
 1     1 (Intercept) -85.0       0      NA        NA           NA  NA                       NA       NA     NA
 2     1 AAEN          0.0424    0 388320.    323385.       95426.  0.0000126             1.20     1.00   TRUE
 3     1 EDML          1.04     43     34.5       33.8     586862.  0.00000174            1.02     0.970  TRUE
 4     1 HFAZ          0.294    18      2.55       2.47    557887.  0.00000185            1.03     0.595  FALSE
 5     1 PPOB         -0.0305   14      4.56       4.51    465461.  0.00000217            1.01     0.778  TRUE
 6     2 (Intercept)  10.7      21      NA        NA           NA  NA                       NA       NA     NA
 7     2 EDM           0.134    31     36.5       22.3    4333882.  0.000000378           1.64     0.955  TRUE
 8     2 EDML         -2.83     28     34.2       22.4    4701988.  0.000000325           1.53     0.955  TRUE
 9     2 FTEN2         0.590    59      4.72       1.94   3550526.  0.000000685           2.43     0.485  TRUE
10     2 FTEN3         0.310    52      4.87       1.83   3144197.  0.000000848           2.67     0.453  TRUE
11     2 FTEN4         0.259    59      4.25       1.97   5012142.  0.000000429           2.15     0.493  TRUE
12     2 HFA2         -0.0264   57      0.959      1.01 11032937.  0.0000000860          0.949    0.0105 FALSE
13     2 HFAZ          0.293    22      0.929      1.02   5940090.  0.000000153           0.907    0.0237 FALSE
```

McFadden R-squared (MCFAR2) are not too high, as expected. The 50, 25 & 75 percentiles for Cook's D (COOKMD, COOKQ1, COOKQ2) are shown. The proportions of Cook's D above2 (COOKP2), above 3 (COOKP3), extreme Beta (BRTAPX), absolute Leverage over 3 (LEVEP3), residuals above 3 (RESIP3) and non-zero covariance matrix terms (MCOVP0) are shown to be very small. The variance inflation factors (svy.vif) are above 3 (FGT3) for most covariates. We interpret this as some collinearity expected (between age terms and between age and anthropometry terms) plus the noise by the complex sample design.

#### Figure S003 Quantile Residual Model Diagnostics (Expanded)

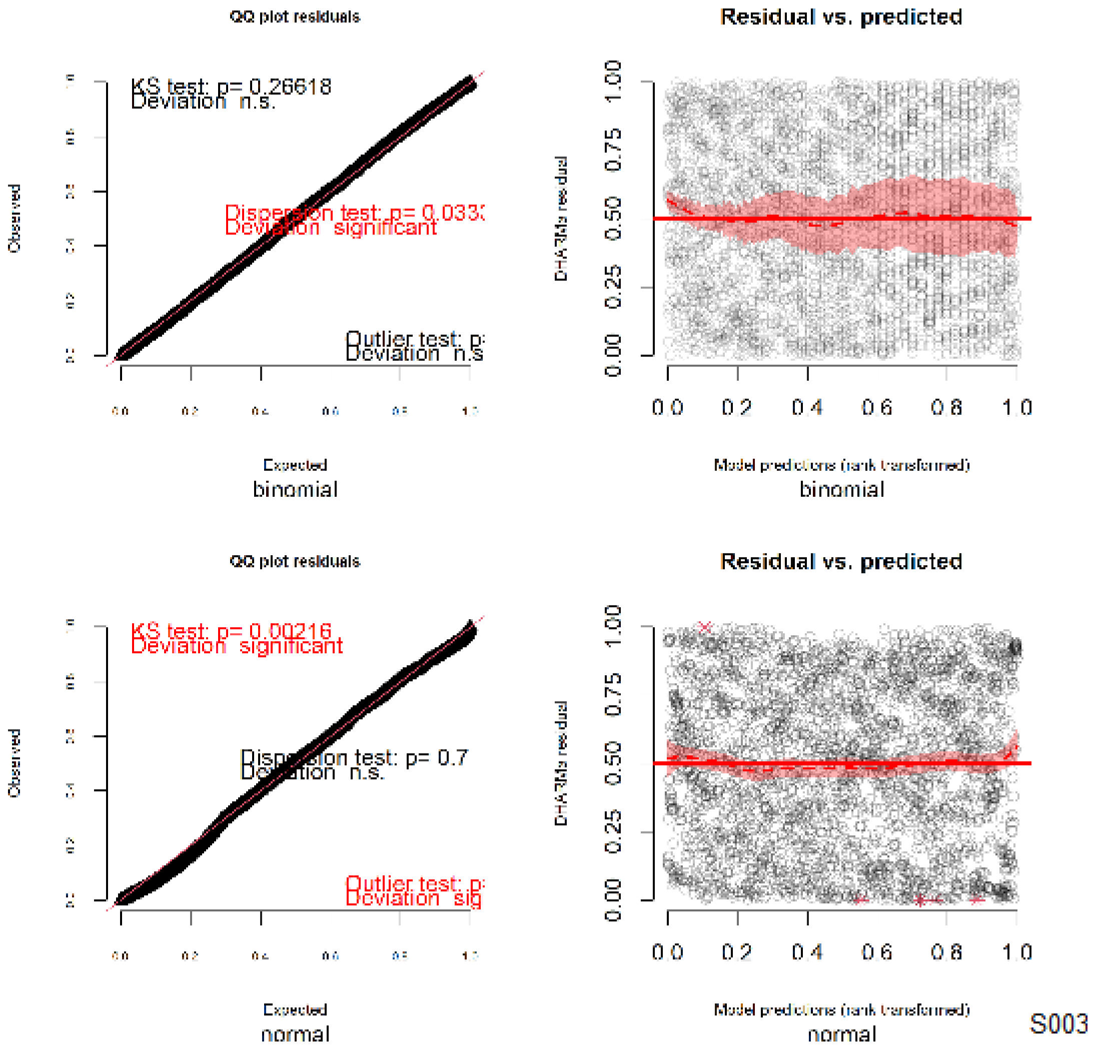

The figure shows the simulated quantile residuals for the model diagnostics. The upper two panels are for the binomial model. The lower two panels are for the normal model. The left-hand panels show the quantile-quantile plot. The right-hand panels show the residuals vs the predictions. Fairly small departures from the straight line are detected for the normal model. A somewhat asymmetric confidence band for the average trend, well within the limits (dashed horizontals) are shown in the right-hand plots.

Our impression is that, while not perfect and not explaining most of the variance, no major departures from the analysis assumptions have been detected, so the final models look reliable.

#### Section S7 Sources of UP Foods

##### Figure G223 Distribution of Food Groups providing NOVA 4 (Weighted)

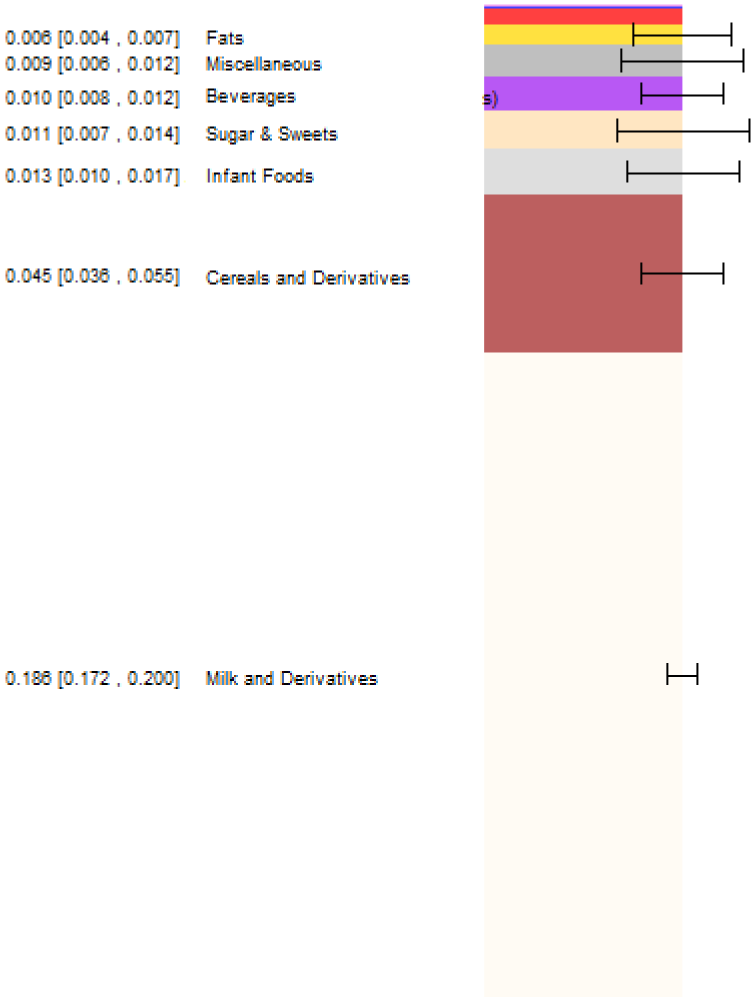

The bars represent the fraction of total energy intake provided by NOVA 4 foods in each food group (each color is a food group, only the top groups are shown). All the bars together add up to the numbers at the bottom of the figure, the fraction of total energy provided by NOVA 4 groups. The numbers on the left represent the weighted fraction estimates, with their 95% confidence intervals within brackets. The error bars on the right side of each bar depict those CI.

41. Peru, Instituto Nacional de Salud, Centro Nacional de Alimentación y Nutrición (CENAN), editor. Tablas Peruanas de Composición de Alimentos. Lima, PE: INS/CENAN; 2017.

42. R Core Team. R: A language and environment for statistical computing [Internet]. Vienna, Austria: R Foundation for Statistical Computing; 2023 [cited 2013 Feb 8]. Available from: http://www.R-project.org

43. Rizopoulos D. bootStepAIC: Bootstrap stepAIC [Internet]. 2022 [cited 2023 Jan 14]. Available from: https://CRAN.R-project.org/package=bootStepAIC

44. Tukey J. Exploratory Data Analysis. Reading, MA: Addison-Wesley; 1977.

45. World Health Organization (WHO). WHO child growth standards: length/height-for-age, weight-for-age, weight-for-length, weight-for-height and body mass index-for-age ; methods and development [Internet]. Geneva: WHO Press; 2006. 312 p. Available from: https://www.who.int/publications-detail-redirect/924154693X
